# COVID-19 vaccine hesitancy among Algerian medical students: a cross-sectional study in five universities

**DOI:** 10.1101/2021.08.29.21261803

**Authors:** Mohamed Amine Kerdoun, Abdellah Hamza Henni, Assia Yamoun, Amine Rahmani, Rym Messaouda Kerdoun, Nazia Elouar

## Abstract

Vaccine hesitancy is a limiting factor in global efforts to contain the current pandemic, wreaking havoc on public health. As today’s students are tomorrow’s doctors, it is critical to understand their attitudes toward the COVID-19 vaccine. To our knowledge, this study was the first national one to look into the attitudes of Algerian medical students toward the SARS-CoV-2 vaccine using an electronic convenience survey.

383 medical students from five Algerian universities were included, with a mean age of 21.02. 85.37% (n=327) of respondents had not taken the COVID-19 vaccine yet and were divided into three groups; the vaccine acceptance group (n=175, 53.51%), the vaccine-hesitant group (n=75, 22.93%), and the vaccine refusal group (n=77, 23.54%).

Gender, age, education level, university, and previous experience with COVID-19 were not significant predictors for vaccine acceptance. The confirmed barriers to the COVID-19 vaccine concern available information, effectiveness, safety, and adverse effects.

This work highlights the need for an educational strategy about the safety and effectiveness of the COVID-19 vaccine. Medical students should be educated about the benefits of vaccination for themselves and their families and friends.

The Vaccine acceptant students’ influence should not be neglected with a possible ambassador role to hesitant and resistant students.

**Graphical abstract:** 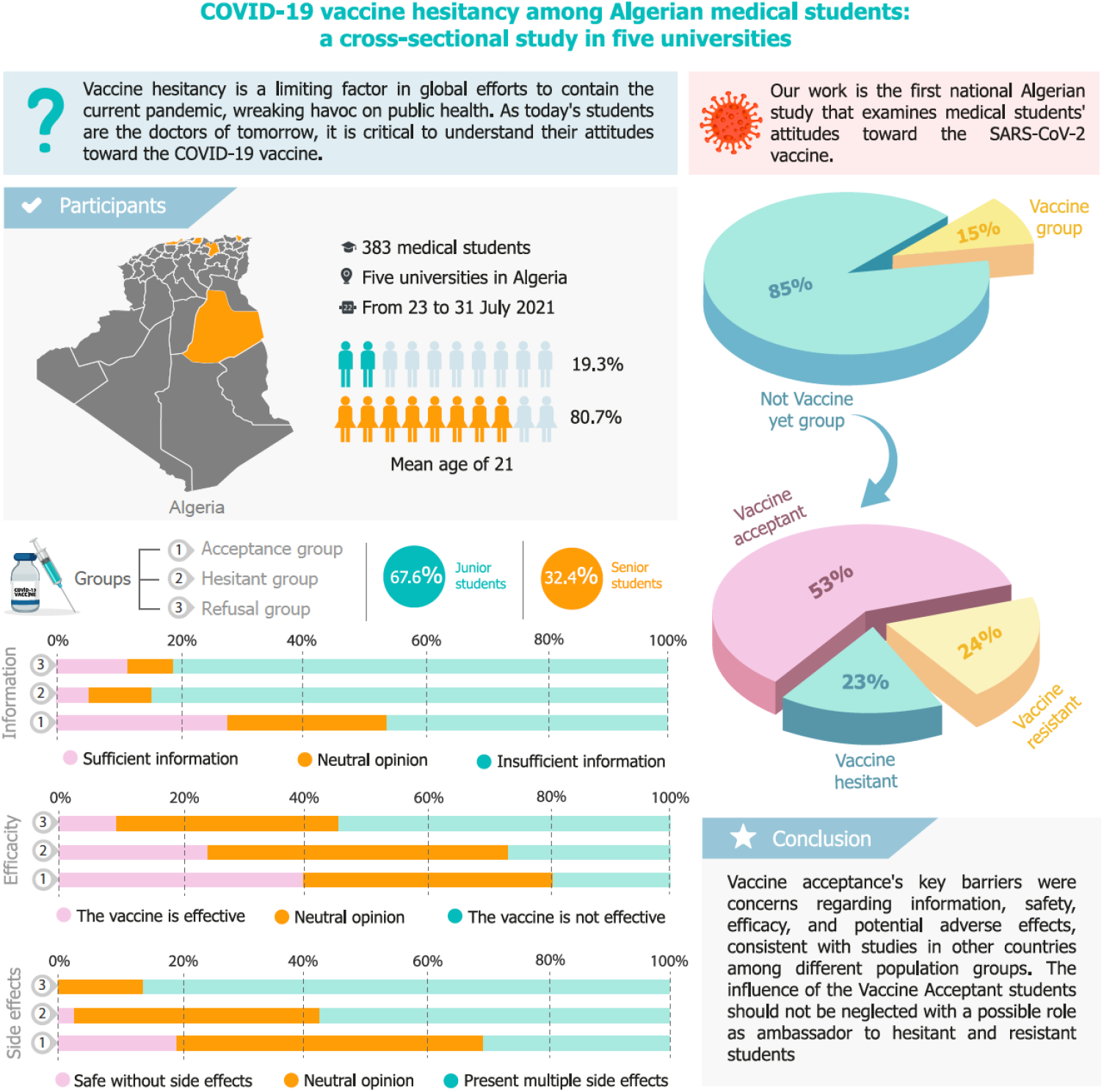

## 1. Introduction

The COVID-19 crisis is a major health pandemic first detected in November 2019 and is a severe acute respiratory disease caused by SARS-CoV-2 [1]. The infection spread quickly in Wuhan, then throughout China and other countries, including Algeria, the largest country of Africa [2, 3]. As of the end of July 2021, 170,189 confirmed cases and 4,219 deaths had been reported in this country.

There are no particular antiviral treatments for COVID-19, and only a few of the already used therapies have demonstrated the ability to decrease mortality in critical patients (e.g., corticosteroids). Additionally, compliance with social distancing and prolonged use of face masks is not guaranteed. Thus, vaccination has been critical in potentially putting an end to the COVID-19 epidemic [4].

Quickly, after the SARS-CoV-2 virus discovery, an extraordinary amount of work has been accomplished. The scientific community has initiated over 300 vaccines projects. 40 vaccines are now being evaluated in clinical trials; several have already received approval from the Food and Drug Administration (FDA) [5] and others are being used in many countries [6, 7].

Although vaccination is a major advance in terms of public health and reduction in mortality, it has been linked from its origins to vaccine hesitancy [8]. Vaccine hesitation refers to behaviors of refusal of certain vaccines or their delivery with delay in a context where vaccination services are functioning [9].

Thus, despite its availability, the public acceptability of the COVID-19 vaccines created in a short time remains uncertain [10]. This hesitation is found worldwide. Concerns about the vaccine’s efficacy or safety, the country of manufacture, antivaccine movements, and the belief in rushed vaccine research were vaccination hesitation causes, in addition to rumors and misinformation [11–13].

In Algeria, the government has approved the Sinopharm and Sinovac Chinese vaccines and the Sputnik Russian vaccine. The national vaccination campaign started by the end of January 2021 for healthcare workers and vulnerable groups. The current local policy targets all the population of the country.

A recent study published in July 2021 described the profile of COVID-19 vaccine acceptability among the Algerian population (49% of participants were healthcare workers) and found that 33.5% agreed to get the vaccination. They determined that two-thirds of Algerians are unlikely to be engaged in COVID-19 vaccine uptake, making them one of the most resistant populations to voluntary vaccination in Arabic countries. [14]. However, the vaccine campaign’s success depends on the vaccination rate [15].

Today’s medical students are tomorrow’s doctors, and they are the future providers of health care and essential influencers of individuals and communities. Additionally, they will be responsible for making vaccination recommendations and counseling vaccine-positive individuals.

At the heart of this COVID-19 pandemic, we need to know and better understand the motivations and hesitations of medical students in the face of vaccination. Documenting the perception and attitude of students towards a vaccination, therefore, appears essential.

The current study targeted medical students from five Algerian universities to explore the level of COVID-19 vaccine hesitancy and determine the factors and barriers that may affect their vaccination decision-making.

## 2. Materials and methods

### 2.1. Study Design and Settings

An observational cross-sectional study was conducted among medical students of five universities in Algeria (Ouargla, Algiers, Annaba, Setif, and Tizi Ouzou) from 23 to 31 July 2021 with a convenience sample and then closed when not receiving any new responses for 24 h.

The questionnaires were electronically distributed via social networks. Volunteer students filled in an anonymous online questionnaire. The inclusion criteria were medical students aged 18 years or older and willing to participate in the study.

### 2.2. Questionnaire

A literature review informed the development of the questionnaire. The questionnaire addressed the following data: personal characteristics, gender, age, residence, college, academic year, and COVID-19 infection. Lastly, vaccination or not, level of acceptance or hesitancy about the COVID-19 vaccine for respondents who do not vaccine yet, beliefs regarding COVID-19 vaccination like side effects, efficacity, and information about the vaccine. The internal consistency was assessed by calculating the Cronbach’s alpha as 0.804. Most of the questions were assigned to be mandatory answer items to avoid incompleteness and missing data.

### 2.3. Statistical analysis of data

Data were extracted from the form to an Excel sheet and statistically analyzed using IBM SPSS software version 23. Descriptive statistics, one-way ANOVA, t-test, and chi-square test were used for the respondents. A significance level of 0.05 was adopted for all statistical analyzes performed.

## 3. Results

A total of 383 medical students [80.67% (n = 302) female and 19.32% (n = 74) male] from five Algerian universities were included, with a mean age of 21.02 (SD=2.15). The students were divided between 67.62% (n = 259) junior and 32.37% (n=124) senior students.

Table 1 summarizes the students’ characteristics and the factors influencing their uptake of the COVID-19 vaccine.

**Table 1.**
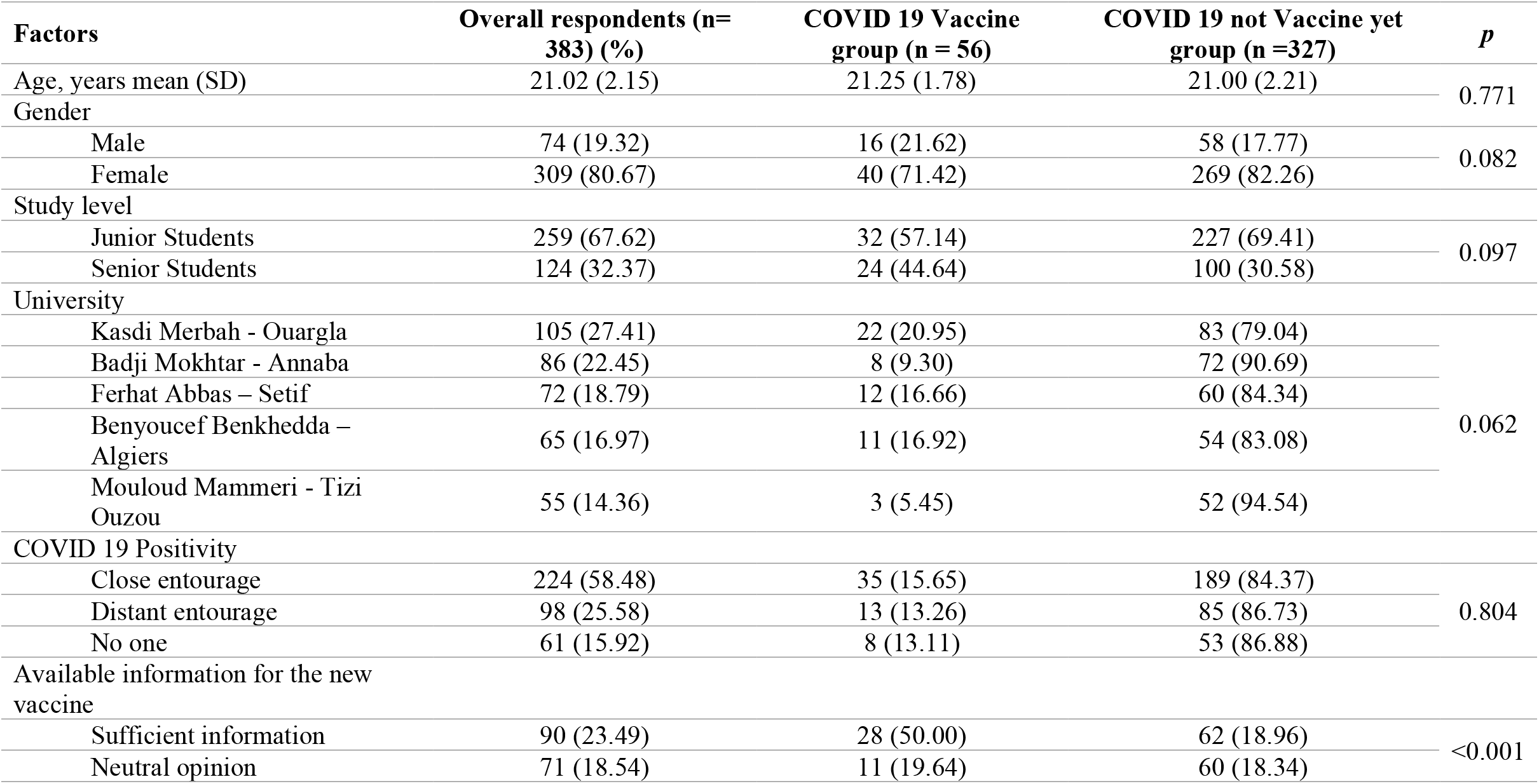
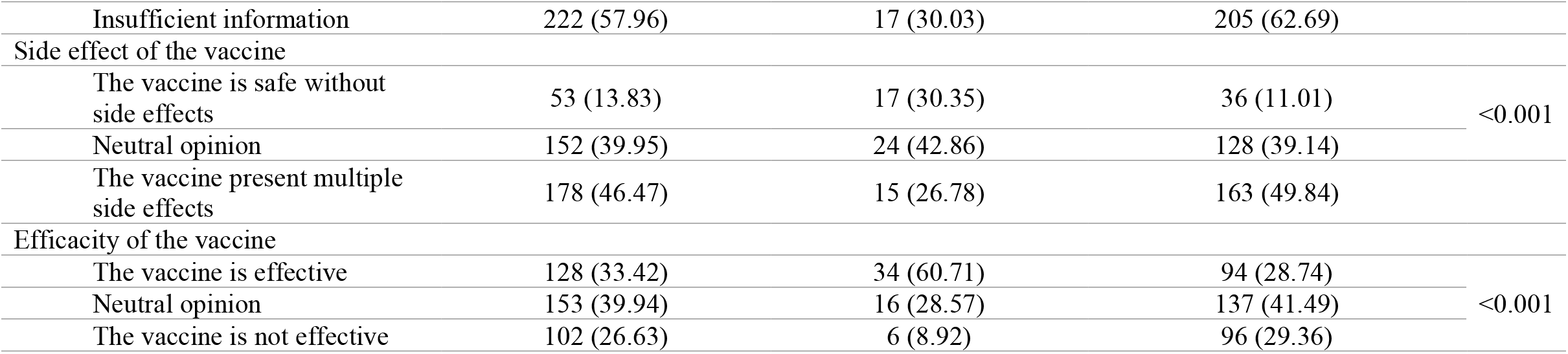
Characteristics of study participants and comparison between vaccine uptake group and non-vaccine group (n=383).

According to Figure 1, 14.62 % (n=56) of respondents have received the SARS-Cov-2 vaccine, while 85.37 % (n=327) have not yet. The non-vaccinated respondents were divided into three groups; the acceptance group (n= 175, 53.51%), the hesitant group (n=75, 22.93%), and the refusal group (n=77, 23.54%).

**Figure 1.**
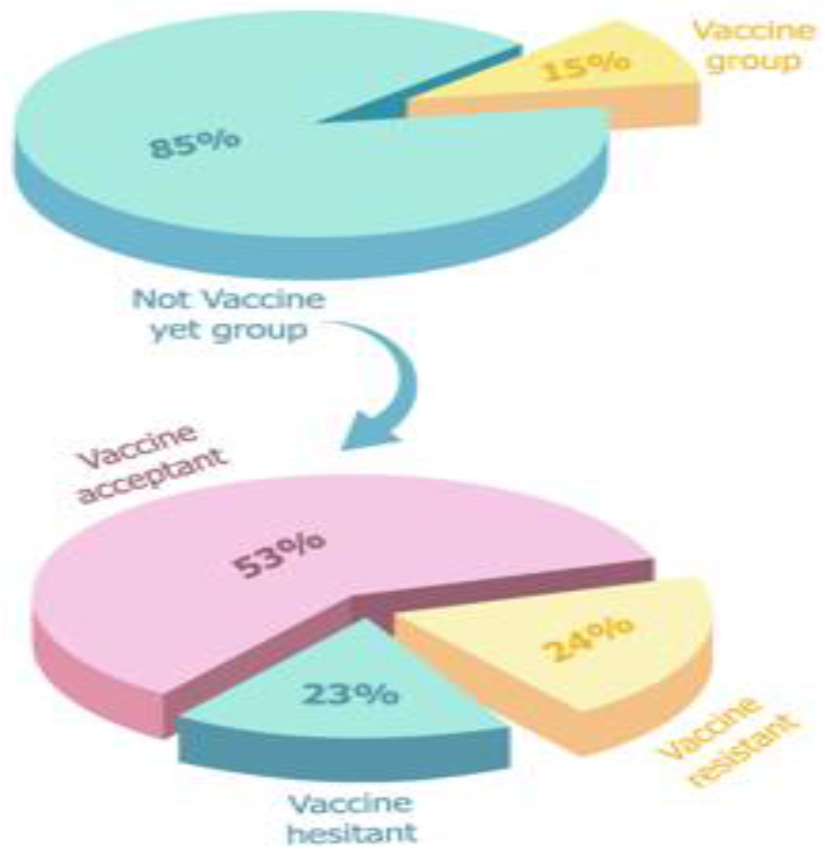
COVID-19 vaccine uptake and acceptance among the respondents.

Demographic variables and experience with COVID-19 infection were not predictive of COVID-19 vaccine uptake, and there was no significant difference concerning age, gender, experience with covid, year of study, or university location.

Confidence in the information, efficacy, and safety of vaccines shows that 57.96% (n=222) of respondents think they have insufficient information, 13.83% (n=53) think that the vaccine is safe, and 33.42 % (n=128) think that the vaccine is inefficient.

Respondents who have received the vaccine feel more informed about the vaccine (50.00% vs. 18.96%), think that the vaccine is effective (60.71% vs. 28.74%), and have no severe side effects (30.35% vs. 11.01%) (p<0.001).

Table 2 shows the characteristics of the non-vaccinated students and factors associated with the COVID-19 vaccine decision.

**Table II.** Characteristics of unvaccinated respondents (n=327).

| **Factors** | **COVID 19 vaccine acceptance group (n=175)** | **COVID 19 vaccine hesitant group (n=75)** | **COVID 19 vaccine refusal group (n=77)** | ***p*** |
| --- | --- | --- | --- | --- |
| Age, years mean (SD) | 21.06 (4.40) | 20.97 (3.40) | 20.99 (6.79) | 0.871 |
| Gender |  |  |  |  |
| Male | 32 (18.29) | 12 (16.00) | 14 (18.18) | 0.902 |
| Female | 143 (81.71) | 63 (84.00) | 63 (81.81) |  |
| Study level |  |  | 1 | 0.671 |
| Junior Student | 118 (51.98) | 53 (23.34) | 56 (24.67) |  |
| Senior Student | 57 (57.00) | 22 (22.00) | 21 (21.00) |  |
| University |  |  |  | 0.144 |
| Kasdi Merbah - Ouargla | 40 (48.19) | 20 (24.09) | 23 (27.71) |  |
| Badji Mokhtar - Annaba | 39 (50.00) | 15 (19.23) | 24 (30.76) |  |
| Ferhat Abbas – Setif | 38 (63.33) | 18 (30.00) | 4 (6.67) |  |
| Benyoucef Benkhedda - Algiers | 30 (55.55) | 10 (18.51) | 14 (25.92) |  |
| Mouloud Mammeri - Tizi Ouzou | 28 (53.84) | 12 (23.07) | 12 (23.07) |  |
| COVID 19 Positivity |  |  |  | 0.162 |
| Close entourage | 109 (57.67) | 41 (21.69) | 39 (20.63) |  |
| Distant entourage | 45 (52.94) | 17 (20.00) | 23 (27.06) |  |
| No one | 21 (39.62) | 17 (32.07) | 15 (28.30) |  |
| Available information for the new vaccine |  |  |  |  |
| Sufficient information | 49 (28.00) | 4 (5.33) | 9 (11.68) | <0.001 |
| Neutral opinion | 46 (26.28) | 8 (10.66) | 6 (13.63) |  |
| Insufficient information | 80 (45.71) | 63 (84.00) | 62 (82.66) |  |
| Side effect of the vaccine |  |  |  |  |
| The vaccine is safe without side effects | 34 (19.42) | 2 (2.66) | 0 (11.01) | <0.001 |
| Neutral opinion | 87 (49.71) | 30 (40.00) | 11 (14.28) |  |
| The vaccine present multiple side effects | 54 (32.57) | 43 (57.33) | 66 (85.71) |  |
| Efficacy of the vaccine |  |  |  |  |
| The vaccine is effective | 69 (39.42) | 18 (24.00) | 7 (9.09) | <0.001 |
| Neutral opinion | 72 (41.14) | 37 (49.33) | 28 (36.36) |  |
| The vaccine is not effective | 34 (19.42) | 20 (26.66) | 42 (54.54) |  |

Medical students are concerned about vaccine hesitancy; only 53.51% are likely to be vaccinated. Being a male and senior student was associated with vaccine acceptance, however, no statistically significant difference has been recorded.

No available information, severe side effects, and inefficacity of vaccine were significantly associated with a higher risk of vaccine hesitancy and refusal (p< 0.001).

## 4. Discussion

### 4.1. Vaccine uptake

The urgency for vaccination is growing by the day, with increasing numbers of cases and reports of the virus variants in the United Kingdom, Brazil, and South Africa spreading to other countries [16]. As of 13 august 2021, 31.1% of the world population has received at least one dose of a COVID-19 vaccine, and 23.4% is fully vaccinated. 4.66 billion doses have been administered globally, and 35.85 million are now administered each day. However, only 1.2% of people in low-income countries have received at least one dose.

Vaccine hesitancy is a limiting factor in global efforts to contain the current pandemic, wreaking havoc on public health and socioeconomic systems [17, 18]. As today students are tomorrow’s doctors; it is critical to understand their attitudes toward the COVID-19 vaccine and work to increase their acceptance to plan an effective post-pandemic strategy. To our knowledge, our study is the first national one to look into the attitudes of Algerian medical students toward the SARS-CoV-2 vaccine.

The results of our research revealed that only 14.62% of students got the COVID-19 vaccine. The proportion of the population that must be vaccinated against COVID-19 to initiate herd immunity is unknown. However, a modeling study determined that vaccines must have an efficacy rate of at least 80% and coverage of at least 75% to end this ongoing epidemic [19].

The low vaccination rates for students can be transposed to the general Algerian population and demonstrate the need for additional efforts to achieve immunization. A recent study conducted in April 2021 to describe the existing profile of potential COVID-19 vaccine acceptance among the Algerian population (healthcare workers constituted 49% of participants), reported that 33.5% accept to receive the vaccine [20].

To increase vaccination coverage, it is necessary to ensure the availability of SARS-Cov-2 vaccines and vaccination promotion. Systematic strategies should be implemented to improve COVID-19 vaccine acceptance and uptake among university students.

According to previous studies, a few combined interventions, such as education and training sessions, easy access to vaccines, and post-vaccination rewards, could significantly increase vaccination uptake [21, 22].

### 4.2. Factors associated with vaccination

This study examined vaccine uptake predictors in great detail. Gender, age, education level, university, and previous experience with COVID-19 were not significant predictors. Our findings indicated that women and junior students were less likely to receive the COVID-19, but these findings were not conclusive (*p*> 0.05).

A global population reports from Kuwait [23], Qatar [24], KSA [25], Jordan [26], and Algeria [20] show that men were more likely to accept the Covid-19 vaccine compared to females [26]. According to a systematic review, women were less likely to be vaccinated during the pandemic [27]. This could be because men engage in riskier behaviors than women [28].

However, past research data showed conflicting results about gender. A global survey including 13,426 individuals in 19 countries with a high COVID-19 burden showed that men were relatively less likely to have a positive attitude towards vaccination than women [29]. Another study showed that women in Russia and Germany had higher acceptance rates of the SARS-CoV-2 vaccines than men [30]. This phenomenon has been named “*the Covid-19 gender paradox*”[31].

Despite their critical role in vaccine promotion and patient counseling, medical students are also concerned about vaccine hesitancy. Of the 327 respondents who had not yet received the vaccine, more than half (53.51%) were willing to take it, a quarter was hesitant (22.93%), and a quarter was resistant (23.54%).

The acceptance rate of our work is lower than a survey of American (75%) [32] and Italian students (86%) [33] conducted in December 2020, comparable to that of French students (57%) in January 2021 [34], and higher than that of Egyptian students (35%) in January 2021 [35]. Also, several studies show that Vaccine hesitancy and refusal among healthcare workers is a veritable concern and mainly originate from the safety and effectiveness of COVID-19 vaccines [36–38].

The respondents who chose not to be vaccinated did not differ significantly in demographic and social characteristics from the rest. However, senior students have more intention of getting vaccinated. According to Barello et al. [33], vaccination attitudes are influenced by students’ knowledge of health issues; students in advanced years are more likely to be vaccinated because of their responsibility to contribute to future waves or peaks.

The majority of participants in this study expressed concerns about vaccine information, effectiveness, safety, and adverse effects. Similar concerns have been raised globally about health workers worldwide [35, 39, 40].

With 57.96%, the primary vaccination barriers identified were a lack of information, and explain why, despite students’ recognition of the COVID-19 vaccine’s importance, they continue to express hesitancy and refusal due to a lack of certainty about vaccination safety and unknown potential adverse effects, in addition to misinformation gleaned from social media [41, 42].

The evidence suggests that it is critical to establish trust in COVID-19 vaccines [43]. Vaccination education during medical school could be improved by using methods based on practical learning, like case-based learning and clinical placements [34]. The positive influence of social networks could also be improved by medical influencers that support the dissemination of scientific insights, including issues related to vaccines and their safety.

Other significant reasons for vaccine refusal and hesitancy include fear of vaccine side effects and doubts about vaccine efficacy. Similarly, Lucia et al. [32], stated that vaccine hesitancy was influenced by concerns about serious side effects and a lack of trustworthy information. These concerns are understandable, given the rapid pace of vaccine development, the novelty of the mRNA technology used in some vaccines, and the public mediatization of vaccine side effects [44, 45]. Many hesitant people about vaccination are expected to accept it after being reassured and provided with credible information about the vaccine’s safety and effectiveness [45].

As future health care workers, medical students should be vaccinated, as vaccination of healthcare professionals and students is critical for preventing health-related infections associated with close contact with high-risk patients [35]. They also should be educated about the benefits of vaccination for themselves and their family and friends [46].

The Vaccine Acceptant students’ influence should not be neglected with a possible ambassador role to hesitant and resistant students. They should be set as an example by encouraging other students and patients to get vaccinated.

## 5. Limitations

This study has some limitations, including an online sample with no random selection and weak generalizability. The percentage of women and junior student were elevated. The study findings are concerned with five universities and thus cannot be generalized. Also, this work was carried out during the third peak in Algeria and may influence COVID-19 vaccine acceptance. Additional research should be based on an equal representation of gender status and include studies across academic disciplines for comparison purposes.

## 6. Conclusion

This is the first study of vaccine hesitancy among medical students in Algeria. The COIVD-19 vaccine acceptance was 57%, with considerably high hesitancy and refusal. COIVD-19 vaccine acceptance’s key barriers were concerns regarding information, safety, efficacy, and potential adverse effects, consistent with studies in other countries among different population groups. The influence of the Vaccine Acceptant students should not be neglected with a possible role as ambassador to hesitant and resistant students.

## Data Availability

The datasets used and/or analyzed during the current study are available from the corresponding author on reasonable request.

## Ethics Statement

All procedures performed in this study involving human participants were following the ethical standards of the University’s Research Ethics Board and with the 1975 Helsinki Declaration.

The institutional review board of Ouargla University has approved this work.

All necessary participant consent has been obtained and the appropriate institutional forms have been archived.

## Acknowledgments

The first author would like to thank all the students who participated in this study, particularly Kasdi Merbah University. “I am very proud to be one of our teachers”.

